# Virtual health care for community management of patients with COVID-19

**DOI:** 10.1101/2020.05.11.20082396

**Authors:** O Hutchings, C Dearing, D Jagers, M Shaw, F Raffan, A Jones, R Taggart, T Sinclair, T Anderson, AG Ritchie

**Affiliations:** Sydney Local Health District

**Keywords:** COVID-19, telemedicine, virtual hospital, community health care, health informatics, health information technology

## Abstract

**Objectives:** To describe the implementation and early experience of virtual health care for community management of patients with COVID-19.

**Design:** observational cohort study.

**Setting:** large Australian metropolitan health service with established virtual health care program and remote patient monitoring capability.

**Participants:** patients with COVID-19 living within the health service who can self-isolate safely, do not require immediate admission to an inpatient setting, have no major active comorbid illness and can be managed at home or other suitable accommodation.

**Main outcome measures:** care escalation rates, including hospital admission.

**Results:** between 11-29 March 2020, 162/173 (93.6%) locally diagnosed patients with COVID-19 were accepted to the virtual health care program, median age 38y (range 11-79). For the 62 patients discharged during this period the median length of stay was 8 days (range 1-17). The peak of 100 prevalent patients equated to approximately 25 patients per Registered Nurse per shift. Patients were contacted a median of 16 times (range 1-30) during this period, with video consultations used 66.3% of the time; 132/162 (81.5%) patients were monitored remotely. Care escalation rates were low: ambulance attendance, 5 (3%); ED attendance, 4 (2.5%); hospital admission, 3 (1.9%). There were no deaths. Conclusions: community-based virtual health care is feasible for managing most patients with COVID-19 and can be rapidly implemented in an urban Australian context for pandemic management. Health services implementing virtual health care should anticipate challenges with rapid technology deployments and provide adequate support to resolve them including strategies supporting consumer use of health information technologies.

## Introduction

Coronavirus disease 2019 (COVID-19) is a respiratory tract infection caused by Severe Acute Respiratory Syndrome Coronavirus 2 (SARS-CoV-2). COVID-19 was declared a global pandemic by the World Health Organisation (WHO) in March 2020(1). The first case was identified in Australia in January 2020 and the situation continues to evolve rapidly.

COVID-19 is a relatively mild disease in the majority of cases; however, approximately 14% develop severe disease requiring hospitalisation and 5% require admission to an Intensive Care Unit(ICU)(2). There is a concern that the Australian health care system will have inadequate access to acute care and ICU beds(3, 4).

Virtual health care (VHC) is a model of health service delivery that substitutes in-person visits with telephone or video consultations and often includes asynchronous data collection from the patient via survey tools or remote monitoring devices. VHC is emerging as a central strategy to manage large numbers of patients affected by the COVID-19 pandemic, as this can maximise the use of limited clinical resources, reduce pressure on acute facilities and reduce the potential for healthcare-associated infection(5). Studies have shown that telehealth has high satisfaction for patients and clinicians, with comparable clinical and service outcomes in chronic disease(6). There have also been case studies of telehealth deployments in other crises, such as in hurricanes Harvey and Irma(7). However, little exists in the literature about the practical issues of fast implementation at scale.

This paper describes the rapid deployment of VHC within a large Australian metropolitan public health service in response to the COVID-19 pandemic.

## Methods

### Setting

Sydney Local Health District (SLHD) is a large metropolitan public health service in New South Wales, Australia, with five hospitals, four large community health centres and over 12,000 employees. SLHD is responsible for the health and wellbeing of approximately 700,000 people living with its boundaries and approximately 1 million visitors who come to the city area each day for work, study and recreation(8).

SLHD has been implementing telehealth and VHC modalities for several years. On 3 February 2020, SLHD commenced operations of RPA Virtual Hospital (**rpa**virtual), Australia’s first metropolitan virtual hospital (9). The service was established as a twelve-month pilot program providing in home and virtual nursing services. The initial cohorts included palliative care patients, adult patients with cystic fibrosis and patients at risk of recurring lower leg wounds. **rpa**virtual is supported by strong operational and clinical governance including the appointment of a General Manager and Clinical Director to oversee its operations and is embedded in the organisational structure of SLHD.

Co-located on the Royal Prince Alfred Hospital campus in Camperdown, NSW, **rpa**virtual comprises of a 24/7 Care Centre that includes technology enabled multi-disciplinary team rooms, handover areas, tracking boards and several Care Pods. The Care Pods provide access to the electronic medical record (EMR), shared care planning and remote monitoring tools. They are equipped with video conferencing and telephones and are staffed by nurses who can remotely monitor multiple patients at once. The facility has medical staff on site and is under the supervision of a Clinical Director and Director of Nursing. In addition to remote consultations, **rpa**virtual has a team of over one hundred community nurses who deliver nursing care in patient homes.

On 5 March 2020, in response to the Australian COVID-19 pandemic, **rpa**virtual began a rapid redesign to provide VHC to patients with COVID-19 managed in the community. The first patients were enrolled on 11 March 2020. In this article we share the early experience using VHC to manage patients with COVID-19 in the community up to and including 29 March 2020.

The Sydney Local Health District Ethics Review Committee reviewed this manuscript and no ethical concerns were raised regarding the study or publication of the results.

### Population

Patients attending COVID-19 Testing Clinics in SLHD in whom SARS-CoV-2 is detected are informed of the result by the local Public Health Unit (PHU) and referred to the **rpa**virtual Care Centre. The Care Centre conducts an initial clinical assessment by telephone to ascertain suitability for VHC. Inclusion criteria and relative exclusion criteria are listed in Table 1. RPA Virtual Hospital Patient Selection Criteria for COVID-19 virtual health care.

**Table 1.** RPA Virtual Hospital Patient Selection Criteria for COVID-19 virtual health care.

|  |
| --- |
| **Inclusion Criteria** |
| • The Public Health Unit is satisfied that the patient can self-isolate safely and understands how to manage self-isolation;<br>• The clinical team is satisfied that it is clinically appropriate to manage the patient at home and a carer can safely care for the patient using appropriate personal protective equipment or the patient can self-care and there is daily monitoring and follow up by a member of the clinical team. |
| **Exclusion Criteria (relative)** |
| • People over 65 years of age with significant comorbidity including, but not limited to cancer, cardiovascular disease, diabetes, heart failure, immunosuppression, stroke, liver disease, renal disease and lung disease;<br>• People under 65 years with one or more of the following comorbidities; lung disease, cardiovascular disease; renal disease (chronic kidney disease stage 5 or requiring renal replacement therapy including renal transplant);<br>• Uncontrolled hypertension;<br>• Residents of Residential Aged Care Facilities (RACFs);<br>• Persons aged <18 years;<br>• Pregnant women. |

Patients with relative exclusion criteria can still be accepted into VHC subject to further discussion with their treating physicians or an Emergency Medicine specialist.

To be able to use the supplied technology and video conferencing, patients require access to a smart phone, tablet or personal computer with internet access and video capability.

Patients unable to self-isolate at home are offered alternative accommodation that is managed by SLHD.

Non-English-speaking patients are accepted with the support of interpreters.

Patients ineligible or unsuitable for VHC are required to be admitted to hospital.

### Model of Care

The **rpa**virtual COVID-19 VHC model is based on early detection of deterioration and managed care escalation for deteriorating patients.

Vital signs, including respiratory rate, oxygen saturation, pulse rate and temperature, are monitored at home. Blood pressure monitoring is not required. Patients are monitored three times per day, including a videoconference with the patient twice every 24 hours, allowing for further assessment of symptoms and signs of deterioration (see Table 2. RPA Virtual Hospital COVID-19 telemedicine patient assessment) based on standard nursing assessment approaches(10, 11).

**Table 2.** RPA Virtual Hospital COVID-19 telemedicine patient assessment.

|  |
| --- |
| **Clinical Progress** |
| 1. Do you feel feverish or have chills?<br>2. Do you have a cough?<br>3. Do you feel short of breath or find it difficult to talk?<br>4. Do you have any other symptoms? |
| **Psychological Screening** |
| 1. How are you managing your home isolation?<br>2. How are you coping generally? |
| **Clinical Assessment** |
| A. Airway;<br>B. Breathing: respiratory distress, respiratory rate, oxygen saturation, other respiratory symptoms;<br>C. Cardiovascular system: appearance, heart rate;<br>D. Disability: alertness, cognition, mental state, mobility;<br>E. Exposure (temperature): temperature;<br>F. Fluid balance: fluid and food intake, gastrointestinal symptoms and losses;<br>G. Glycaemic control (if diabetic): blood glucose readings; |

Vital signs are recorded electronically in the EMR and tracked against a standardised early warning system, known as ‘Between The Flags’, using the Standard Adult General Observation chart criteria(12, 13).

Clinical escalation of deteriorating patients is a managed transfer to the local Emergency Department (ED), via ambulance if clinically indicated. The ambulance service is notified of the patient’s infectious status and advised to call the receiving ED prior to arrival. In the event of escalation, the Care Centre will also notify hospital executive and the local PHU.

A communication escalation pathway is activated if a patient cannot be reached by the Care Centre. If a patient cannot be contacted after one phone call, an SMS is sent. If the patient does not call back within one hour, a second SMS is sent expressing concern for their welfare. If the patient does not answer a third phone call, the NSW Police are contacted to conduct a welfare check.

Medical Officers at **rpa**virtual are consulted by referral from the Care Centre staff to discuss patient deterioration, escalation decision-making, medical certification and prescribing medications, although medication management is not a component of the model of care.

Discharge from VHC is in accordance with the ‘release from isolation’ criteria in the Coronavirus Disease 2019 (COVID-19) CDNA National Guidelines for Public Health Units(14):

- The person has been afebrile for the previous 48 hours
- Resolution of the acute illness for the previous 24 hours
- Be at least 7 days after the onset of the acute illness
- PCR negative on at least two consecutive respiratory specimens collected 24 hours apart after the acute illness has resolved

Patients meeting the first three criteria are referred to a COVID-19 Testing Clinic for repeat testing. After negative results are confirmed, the patient is discharged from VHC and referred to their GP for ongoing care.

### Patient Experience

The use of technology by patients and carers is central to care in **rpa**virtual.

Once accepted in VHC, new patients are provided with a welcome pack containing the following items:

- A welcome letter and videoconferencing instructions
- Pulse oximeter and instructions
- Temperature monitoring device and instructions
- NSW Government COVID-19 factsheets
- A patient responsibilities pamphlet

Personal protective equipment (PPE) is provided immediately following COVID-19 testing and a resupply is delivered with the welcome pack.

The welcome pack is delivered by a “Flying Squad” of health informatics staff wearing appropriate PPE following completion of a home visit risk assessment. The **rpa**virtual Flying Squad instruct patients to wear personal protective equipment when answering the door and notify patients when they are 5 minutes away.

Patients interact with the **rpa**virtual team through video consultations and via a 1800 free call telephone service. Calls are scheduled with the patient to collect their observations. Patients are advised to call the **rpa**virtual Call Centre or an ambulance if they deteriorate.

### Remote Monitoring Technology

Remote monitoring devices were evaluated to collect patient generated health data from patients with COVID-19 according to defined criteria (see Table 3. RPA Virtual Hospital Patient Remote Monitoring Technology Assessment Criteria).

**Table 3.** RPA Virtual Hospital Patient Remote Monitoring Technology Assessment Criteria.

| Technology Evaluation Criteria<sup>^</sup> |
| --- |
| 1. Capable of measuring desired clinical observations (heart rate, oxygen saturation, temperature and respiratory rate);<br>2. Capable of remote data collection and transmission (or able to be read by the patient);<br>3. Usability for RPA Virtual Hospital medical team;<br>4. Usability for patients in the RPA Virtual Hospital COVID-19 patient context;<br>5. Training and support requirements for SLHD health informatics staff<br>6. Listed on the Therapeutic Goods Administration's Australian Register of Therapeutic Goods, or able to be fast tracked (e.g. CE or FDA marked);<br>7. Supply chain availability to meet the scale, speed and demand of COVID-19 response;<br>8. Cost;<br>9. Compliance with cyber-security, data security and privacy and infection control requirements. |

<sup>^</sup>Not in ranked order.

No suitable single device was identified that met all vital sign monitoring requirements against the criteria above.

Two devices were selected to use in the COVID-19 program, a pulse oximeter and temperature patch. No reliable device for measurement of respiratory rate was identified and this is measured during the face-to-face videoconference.

A wireless pulse oximeter (iHealth^®^Air pulse oximeter PO3M, iHealth Labs, Inc., Sunnyvale, CA, USA), provides peripheral oxygen saturation and pulse rate measurements. Pulse oximeters are only used by one patient; they are not reused as they cannot be adequately disinfected.

A single-use, wearable temperature monitor (Temp°Traq^®^ Clinical, Blue Spark Technologies, Inc., Westlake, OH, USA) is self-applied in the axilla and provides continuous temperature monitoring. The device feeds continuously into a web-based dashboard, providing the Care Centre with a summary view of all patients. Each patch lasts for 72 hours and each patient is provided with three patches to cover the first 9-11 days of isolation.

Both devices have Bluetooth® connectivity, but only the temperature monitor requires connection to a compatible Apple® or Android™ mobile device to be read; the pulse oximeter can be read directly from the device. Both devices are FDA approved, CE marked and registered with the TGA or able to be fast-tracked, with limited evaluation in clinical settings(15, 16).

The Flying Squad prepares devices for delivery, including pre-charging the pulse oximeters and registering all three temperature monitoring patches to the specific patient. After delivery, they contact the patient by telephone and assist them to download and set up the required mobile application, allowing temperature readings to feed continuously into a web-based dashboard. The Care Centre has a summary view of all prevalent monitored patients with colour-coded parameters to identify patients with an abnormal temperature or patches that are not functioning normally (see Figure 1).

**Figure 1.**
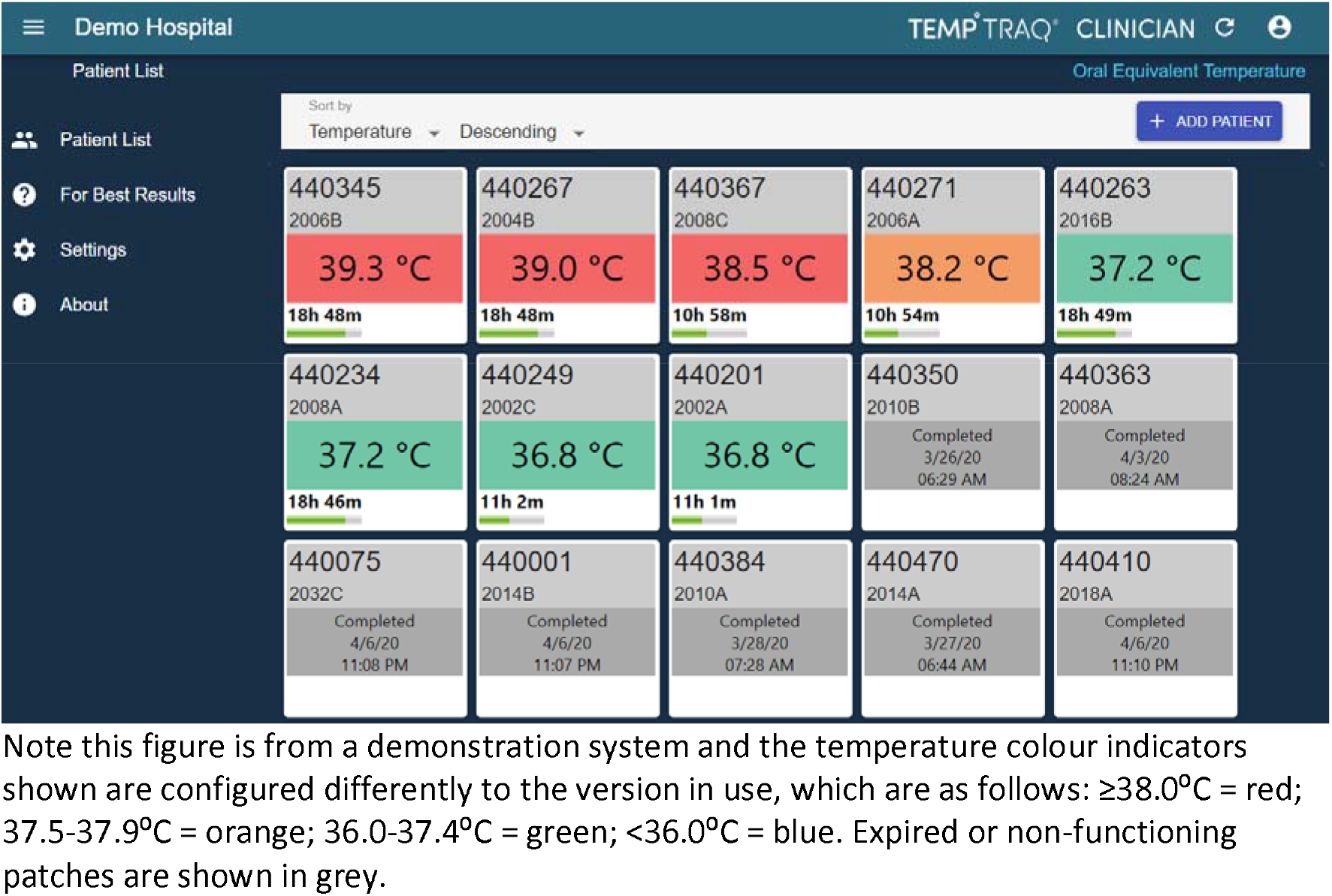
Screen capture of the temperature monitoring portal.

Patient-reported observations are documented in the electronic medical record (Cerner Millennium, Cerner Corporation, Kansas City, MO, USA) in purpose-built sections to allow them to be differentiated from other methods of recording vital signs.

### Video Consultations

Video consultations were implemented to sight the patient, confirm vital signs collected from wearables devices and to estimate the respiratory rate.

The **rpa**virtual Care Pods are fitted with high-definition web cameras and Bluetooth enabled headsets to allow the patient to see and hear the nurse. The Care Pods are configured to enable conversation with the patient to be kept private. There are pull up backgrounds and feature walls behind each nurse to minimise distractions on the video.

A commercial web-based video conferencing solution (Pexip^®^ AS, Oslo, Norway) is used for remote video consultations. This platform works on both desktop and mobile devices and is endorsed by eHealth NSW.

Patients call in to video consultations using their own computer or mobile device.

## Results

Between 11 and 29 March 2020 (inclusive) 5821 people were tested for SARS-CoV-2 at SLHD COVID-19 Testing Clinics and infection was detected in 173 people. Of these, 162 (93.6%) were accepted to the **rpa**virtual program with the remainder admitted to hospital or referred to another health service. The median age of admissions was 38 years (range of 11-79); three patients <18 years and two pregnant women were accepted for VHC.

The median admissions per day was 6 (range 1-30). A total of 62 patients were discharged during this period with a median length of stay of 8 days (range 1-17 days). At the end of the period, there were 100 prevalent patients receiving VHC.

The Care Centre commenced operations with 4 full-time equivalent (FTE) Registered Nurses and gradually increased to 9.5 FTE at the end of the period. This equated to a ratio of approximately 25 patients per Registered Nurse per shift.

Patients were contacted a total of 2865 times, with a median of 16 contacts per patient (range 1-50). Video consultations (1902, 66.3%) comprised most patient contacts with telephone consultations (688, 24.0%) making up most of the remainder. The ratio of telephone consultations to video consultations was 1:2.8. The median duration of each contact was 8.5 minutes (IQR 5-15) for telephone consultations and 15 minutes (IQR 13-15) for video consultations.

Between 18 and 29 March 2020 inclusive, the **rpa**virtual Flying Squad delivered welcome packs with remote monitoring to 132/162 (81.5%) patients.

Ambulances were called to attend five patients, and four were transferred to the ED for assessment. Three patients were subsequently admitted, and one was discharged home to continue with VHC. There were no deaths and no referrals to the Police for welfare checks. However, one person was subject to a public health order for failing to adhere to self-isolation requirements.

Detailed patient outcomes will be reported separately.

## Discussion

We describe the rapid development of a model of care and technology model to deliver VHC for community management of patients with COVID-19. We found that a model of care that excludes high-risk patients with COVID-19 was able to accept the majority of locally identified positive cases. Using remote clinical appraisal, supported by home monitoring of clinical observations, there were low observed rates of deterioration requiring escalation and no patient deaths. The program was also able to accept pregnant women and paediatric patients, expanding the range of patients who may be suitable for VHC for COVID-19 as an alternative to hospital inpatient admission.

A uniform model of care for all patients, regardless of care needs or risk of deterioration, may not be appropriate or necessary. Supply chain delays may also impact rapid procurement of adequate quantities of the necessary technology to remote monitoring, which may be further compounded by limitations on the ability of the Flying Squad to deploy devices to patients. Risk stratification at admission with enhanced monitoring of those more likely to deteriorate is being considered, although there is little empirical evidence to guide that strategy(17). Criteria Led Discharge, a process in which discharge is determined from pre-defined clinical parameters, is also being explored with the aim of expediting discharge from the service(18).

Mental health and well-being are issues requiring further consideration in the provision of VHC to patients with COVID-19. Quarantine and self-isolation for the purpose of controlling infectious diseases is associated with negative psychological effects including post-traumatic stress symptoms, confusion, and anger, with potential to be long-lasting(19). Two psychological assessment screening questions were included in the model to assist with recognising mental health concerns. The psychological impact of self-isolation in one’s own home may be different to that of individuals placed in mandatory quarantine in hotels following arrival from overseas. This is an issue requiring further exploration and consideration is being made to provide access to Social Workers, Psychologists and Psychiatrists through VHC.

Rapid changes to the EMR were required to support VHC for patients with COVID-19, however substantial opportunities for improvement remain. EMRs can be useful tools for rapid deployment of standardised processes, including in response to the COVID-19 pandemic(20). To support the redesign of **rpa**virtual for care of patients with COVID-19, new locations and patient lists were created in the EMR. Access to the electronic between the flags charts for detecting deterioration, previously only used in the inpatient setting, was extended to all Care Centre staff for use in the community. Clinical documentation templates, simple reports and modifications to the results flowsheet to allow recording of remote clinical observations were designed and implemented. However, communication systems are managed separately from the EMR, with separate systems used for messaging, phone calls and videoconferencing. Clinical observations from the patient’s devices are captured separately and manually entered into the EMR. A comprehensive and more integrated suite of digital health care tools would reduce fragmentation of information systems and workflows and improve service delivery through automation of manual processes. There is also a desire for better tools to manage patient flow and overall population management.

A range of technical and operational issues are to be expected with rapid implementation of video consultations and this was true of our experience(6). The model of care required three patient contacts per day with a minimum of two to be conducted via video. This was to ensure adequate surveillance of visual signs of deterioration and to measure the respiratory rate directly. The reported experience of patients with COVID-19 indicated that the most likely time of deterioration was around day 7 after symptom onset, which was the median time of hospitalisation in those who developed an associated pneumonia(21). Initially, patients were scheduled for 10-minute appointments for their video and telehealth consultations. When a problem with video consultation was encountered that staff could not resolve quickly, they would revert to a telephone consultation instead. This is an example of workaround: a common phenomenon when using health information technology that is driven by the need to resolve conflicting goals in a timely fashion(22). In this case, the workaround was driven by the need to adhere to the schedule of appointments rather than delay to resolve the issue. Improving the user experience for staff and patients will require both technical and workflow changes, with fixed appointment slots abandoned. The optimal approach to education and training in VHC, particularly with a rapid increase in staff redeployed from other areas of the business, in the context of high service demands, requires further consideration.

The need for enhanced support for patients to use health information technologies was identified early in the implementation. The model of care is dependent upon patients or their carers being able to use medical technology (pulse oximeter and temperature patch) and digital health applications (for temperature monitoring and video consultations). The use of health information technology by consumers, known as consumer health informatics, has a strong focus on usability and accessibility(23, 24). It was quickly recognised that patients enrolled in VHC could not easily download the temperature monitoring application and connect it with the patches using the instructions provided in the welcome pack alone. The Flying Squad delivering welcome packs, comprising a multidisciplinary health informatics team, were ideally suited to providing additional support. A process was established to contact patients shortly after delivery of the welcome pack and support them in achieving the goal of transmitting temperature readings to the cloud-based monitoring panel. If consumer health technology is to be relied upon for healthcare delivery, health services will require a strategy to support it. Whilst this has been explored in the chronic disease setting, in which there is an opportunity for patients to attend a health service for in-person support, this cannot be done during an infectious pandemic(25). Managed mobile health care platforms, in which patients are provided with a tablet computer with all relevant applications pre-installed and connected to relevant peripherals, may simplify and improve the patient experience and this approach has been used by **rpa**virtual with other patient cohorts. The role of health informaticians in supporting consumer health informatics is a little explored aspect of these emerging roles meriting further exploration.

In summary, community-based VHC is a feasible and safe approach to managing less severe cases of COVID-19 and can be rapidly implemented in the Australian context for pandemic management with strong operational and clinical governance including integration with clinical specialists. Health services implementing VHC should anticipate challenges with rapid technology implementations and provide adequate support to resolve them including strategies to support consumer use of health information technologies.

## Data Availability

Data from this study may be made available on request to the authors.

